# Comparing the diagnostic and clinical utility of WGS and WES with standard genetic testing (SGT) in children with suspected genetic diseases: A systematic review and meta-analysis

**DOI:** 10.1101/2023.07.17.23292722

**Authors:** Kimberley M.B. Tirrell, Helen C. O’Neill

## Abstract

3.0

**Importance:** Rare genetic diseases are one of the leading causes of infant mortality worldwide. Whole-genome sequencing (WGS) and whole-exome sequencing (WES) are relatively new techniques for diagnosing genetic diseases, that classic newborn screening (NBS) fails to detect.

**Objective:** To systematically assess the diagnostic and clinical utility of WGS and WES, compared to standard genetic testing (SGT), in children with suspected genetic diseases, and discuss its impact on the expansion of NBS.

**Data Sources:** EMBASE, MEDLINE, PubMed, Scopus, Web of Science, Cochrane Central Register of Controlled Trials, and references of included full-text articles were searched until 21^st^ October 2021.

**Study Selection:** Studies reporting the diagnostic yield or rate of change of management for WGS and/or WES were included. The meta-analysis included 43 of the original 1768 identified articles (2%).

**Data Extraction and Synthesis:** Data extraction followed the Preferred Reporting Items for Systematic Reviews and Meta-analyses reporting guideline. The quality of included papers was assessed using QUADAS-2, and a meta-analysis was performed using a random-effects model to create pooled proportions and a pooled odds ratio.

**Main Outcome(s) and Measure(s):** Diagnostic utility, as determined by the diagnostic yield, which is defined as P/LP variants with strong or moderate associations with the presenting clinical phenotype of the affected patient, and that were reported to the patient’s clinician. Clinical utility as defined by any change in clinical management (medically or surgically), determined through clinician questionnaires or Electronic Health Record reviews.

**Results:** A total of 43 studies were included, comprising 6168 children. The pooled diagnostic utility of WES (0.40, 95% CI 0.34-0.45, *I^2^*=90%), was qualitatively greater than WGS (0.34, 95% CI 0.29-0.39, *I^2^*=79%), and SGT (0.19, 95% CI 0.13-0.25, *I^2^*=64%). The pooled clinical utility of WGS (0.74, 95% CI 0.56-0.89, *I^2^*=93%), was qualitatively greater than WES (0.72, 95% CI 0.61-0.81, *I^2^*=86%), while both were qualitatively greater than SGT (0.69, 95% CI 0.38-0.94).

**Conclusions and Relevance:** Our evidence suggests that WGS/WES should be considered the first-line test for genetic diseases. There is reason to believe that WGS and WES should be included as part of NBS, however, more studies are required to assess the cost-effectiveness of this approach.

## 4.0 Introduction

Genetic disorders, including monogenic diseases and chromosomal abnormalities, are one of the leading causes of infant mortality, particularly among those admitted to the neonatal and paediatric intensive care units.(^1, 2^) An estimated 400 million people worldwide are thought to suffer from a rare disease of which 80-85% are believed to have genetic origins. Approximately half of those affected by a rare disease are children, with 30% not surviving past their fifth birthday.(^3, 4^) It has been estimated that around 50% of patients with a genetic disorder are never diagnosed.(^5^) Although individually each genetic disease is rare, when combined, the estimated 6000-7000 diseases, are common and contribute significantly to infant morbidity, mortality, and healthcare costs.(^6, 7^)

Disease progression of genetic disorders within children can be rapid, and without early etiological diagnosis, intervention and management decisions made are uninformed and often ineffective, exacerbate symptoms or cause adverse effects, and lead to delays in starting appropriate treatment.(^8–10^) Therefore, a quick, accurate diagnosis for children is vital to improve outcomes, and reduce morbidity and mortality.(^10^)

Attaining a diagnosis for every child with a suspected genetic disease remains a significant challenge, due to the genetic and phenotypic variation of such diseases. Many countries have implemented newborn screening (NBS) programmes in an effort to reduce infant mortality associated with rare diseases, however, these programmes, where implemented, fail to recognise and screen for many rare genetic diseases. Although the WHO have published guidelines for the inclusion of a condition in NBS programmes, there remains to be large disparities between the conditions screened for in many countries and their individual states.(^11–14^) Any abnormalities detected during screening can provide an early indication of a rare disease, however, any rare diseases not detectable through analytes, such as some rare genetic diseases, cannot be screened for. Expanding the list of conditions for NBS to include other rare diseases, not detected through analytes, would ensure a broader range of conditions can be rapidly diagnosed and treated, improving outcomes and reducing infant morbidity and mortality.

Next-generation sequencing (NGS) technologies have rapidly advanced in recent years, and have shown great promise of new diagnostic potentials, due to their genetic and phenotypic approach. Whole-genome sequencing (WGS) and whole-exome sequencing (WES) allow for simultaneous analysis of numerous genes associated with genetic disorders, an approach that is not currently utilised within NBS.(^15^) The speed at which these approaches can analyse genomic data and identify pathogenic/likely pathogenic (P/LP) variants, makes them prime candidates for the expansion of NBS, due to their capabilities of rapid, early diagnoses of additional disorders that are not currently screened for, and would benefit from early detection and subsequent treatment. Here, we report a literature review and meta-analysis of the diagnostic and clinical utility of WGS and WES, compared with standard genetic testing (SGT), in children (≤18 years) with suspected genetic diseases, and discuss the impact this has on the expansion of the NBS programme through WGS and WES.

## 5.0 Methods

### 5.1 Data sources and record identification

On 21^st^ October 2021, we searched MEDLINE, EMBASE, Cochrane Central Register of Controlled Trials, PubMed, Scopus and Web of Science with the MeSH terms (“Infant” or “Infant, Newborn” or “Child”), and (“Whole Genome Sequencing” or “Whole Exome Sequencing” or “Genetic Testing” or “High-Throughput Nucleotide Sequencing”), and (“Critical Illness” or “Intensive Care Units” or “Intensive Care, Neonatal” or “Intensive Care Units, Pediatric” or “Intensive Care Units, Neonatal” or “Critical Care”), and relevant key terms. We manually searched the references of included papers for any missed eligible papers. There were no date, language, or literature type restrictions on searches. Papers identified through database searches were imported into EndNote X9 (Clarivate Analytics, Boston, MA) for duplication removal, title and abstract screening, and full-text review. Full search strategies are available within the appendix (**Appendix II**).

### 5.2 Inclusion criteria and study eligibility

Studies that assessed the diagnostic utility or clinical utility (proportion of patients tested who had a change in clinical management upon receiving a diagnosis) of WGS and/or WES were eligible. Studies containing cohorts with specific disease types or clinical presentations, rather than a broad range of potential genetic diseases, probands over 18 years of age, already diagnosed or containing expired probands were excluded. Case reports, meeting/conference abstracts, and studies where full-texts were not available in English were also excluded. The review was performed according to the Preferred Reporting Items for Systematic Reviews and Meta-analyses statement (**Appendix I, Table 1**).(^16^)

### 5.3 Data extraction

Data extracted comprised of (1) the methodological information of the studies, including: first author, year of publication, objectives, sequencing method, sample size, and study country, (2) patient demographics, including: age of participants and rate of consanguinity, and (3) reported study outcomes, including: diagnostic yield, change in management, incidence of VUS, incidental findings, incidence of de novo variants, and turnaround time was extracted manually. Data was reviewed for completeness and accuracy by two authors with any disparities resolved by discussion and consensus. The PICOTS typology of the criteria for inclusion of studies in quantitative analyses was:

#### Patients

Data extraction was limited to critically ill children (aged less than 18 years) with a suspected genetic disease.

#### Intervention

WGS and/or WES

#### Comparator

Participants tested by WGS, WES and SGT were grouped and compared. SGT was treated as the Reference Standard.

#### Outcomes

Diagnostic utility and clinical utility. Diagnostic utility was determined by the diagnostic yield, which is defined as P/LP variants with strong or moderate associations with the presenting clinical phenotype of the affected patient, and that were reported to the patient’s clinician.(^17^) Clinical utility was defined as any change in clinical management (medically or surgically) as determined through clinician questionnaires or Electronic Health Record (EHR) reviews. Incidental findings and variants of uncertain significance, where available, were also extracted.(^18^)

#### Timing

Where more than one paper reported results from the same study cohort, we extracted the most recent data for diagnostic and clinical utility.

#### Settings

There were no setting restrictions.

### 5.4 Quality assessment

Quality assessment involved evaluating the risk of bias for each included study using the QUADAS-2 tool, a validated tool for assessing the risk of bias in primary diagnostic accuracy studies.(^19^) The QUADAS-2 tool enables the classification of studies into low risk, high risk or unclear risk based on the following domains: patient selection (bias as a result of the selection of participants and representativeness of the sample), index test (bias as a result of the conduction and interpretation of the index test), reference standard (bias as a result of the conduction and interpretation of the reference standard), and flow and timing (bias as a result of the time interval and any interventions between the index test and reference standard). Applicability of studies was also evaluated for the first three domains in each study and judged as “yes, no, or unclear”, indicating a low, high, and unclear risk of bias, respectively.

### 5.5 Statistical analysis

Meta-analysis was conducted using the ‘metaprop’ and ‘metan’ commands in Stata version 15.(^20^) We transformed proportions from individual studies by stabilizing the between-study variance, using the Freeman-Tukey double arcsine transformation procedure, before computing the weighted overall pooled estimates, using the DerSimonian-Laird random-effect model, with an estimate of heterogeneity being taken from the inverse-variance fixed-effect model.(^21^) 95% confidence intervals are based on exact binomial procedures (Clopper-Pearson interval). The chi-squared test was used to assess between-study heterogeneity, with I^2^ statistic values of 25%, 50%, and 75% interpreted as low, moderate, and severe heterogeneity, respectively.(^22^) Forest plots were used to summarize the individual study and pooled group meta-analysis statistics.

### 6.0 Results

### 6.1 Literature search results

WGS and WES are fast becoming commonplace methods for the diagnosis of genetic diseases. We compared the diagnostic and clinical utility of WGS and WES with that of SGT, including chromosomal microarray (CMA), Sanger sequencing, single-gene testing, panel testing, methylation studies, NBS, and others, as the standard of care for children with suspected genetic diseases. A total of 2635 records were identified through searches for studies assessing the use of WGS and WES in children with a wide range of suspected genetic diseases. Thirty-six of these records, comprising 5681 children, met the eligibility criteria. A further seven records were identified through manual searching of included records’ reference lists, bringing the total number of eligible, included records to forty-three, comprising 6168 children.(^10, 23–64^) Of the forty-three included studies, thirty-eight were case studies; five were randomized controlled trials.(^35, 38, 41, 47, 60^) The process and outcome of the literature search are presented in detail in **Figure 1**.

**Figure 1:**
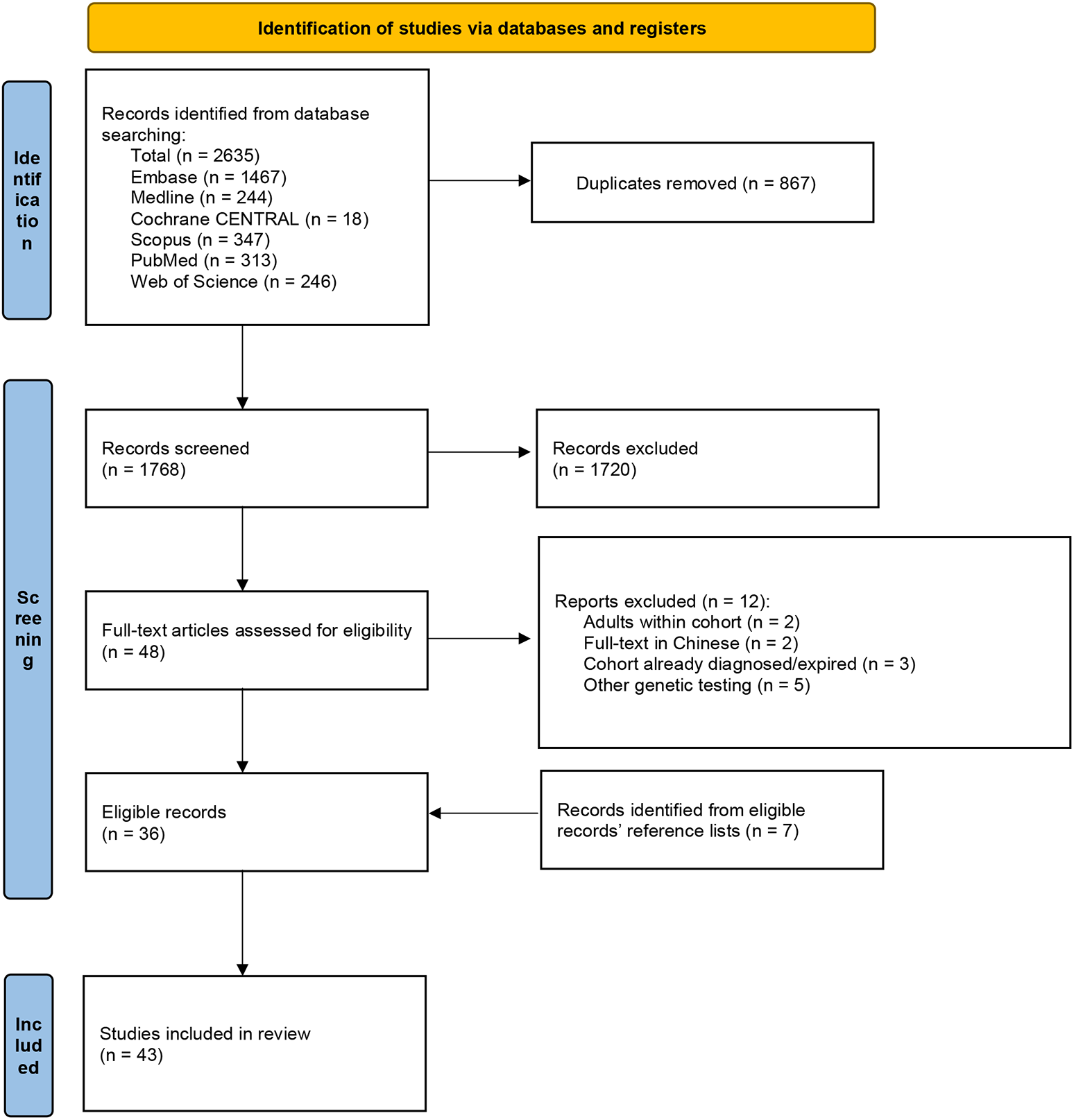
Prisma (Preferred Reporting Items for Systematic Reviews and Meta-Analyses) flow diagram for meta-analysis of diagnostic and clinical utility of WGS and WES.

### 6.2 Meta-analysis results

Of the 43 included studies, 15/43 (35%) looked specifically at WGS, while 25/43 (53%) investigated WES. The characteristics of all forty-three included studies can be found in **Appendix I**, **Table 2**. The pooled diagnostic utility of WES was 0.40 (95% CI 0.34-0.45, 27 studies, 4238 children, *I^2^*=90%), which was qualitatively greater than WGS (0.34, 95% CI 0.29-0.39, 17 studies, 1817 children, *I^2^*=79%), and SGT (0.19, 95% CI 0.13-0.25, 6 studies, 669 children, *I^2^*=64%) (Figure 2). The pooled clinical utility of WGS was 0.74 (95% CI 0.56-0.89, 13 studies, 467 children, *I^2^*=93%), which was qualitatively greater than WES (0.72, 95% CI 0.61-0.81, 18 studies, 648 children, *I^2^*=86%), and SGT (0.69, 95% CI 0.38-0.94, 2 studies, 12 children) (Figure 3). *I^2^* could not be assessed for SGT due to the small sample size of studies. Severe heterogeneity (*I^2^*>75%) within WGS and WES groups precluded statistical comparisons. Among studies that provided complete data for the diagnostic utility of WGS or WES and SGT, the pooled odds of diagnosis were 2.93 times greater for WGS/WES (P<0.01) (Figure 4). 31/43 (72%) studies reported the heritability of detected variants, these included P/LP variants, variants deemed to be an incidental finding, and VUS. A total of 596/1381 (43%) were de novo variants. Some studies opted out of reporting incidental findings, while others only returned incidental findings to patients and families who had consented. Of the eighteen studies opting to report incidental findings, a total of 66/1221 (5%) participants received such findings.

**Figure 2:**
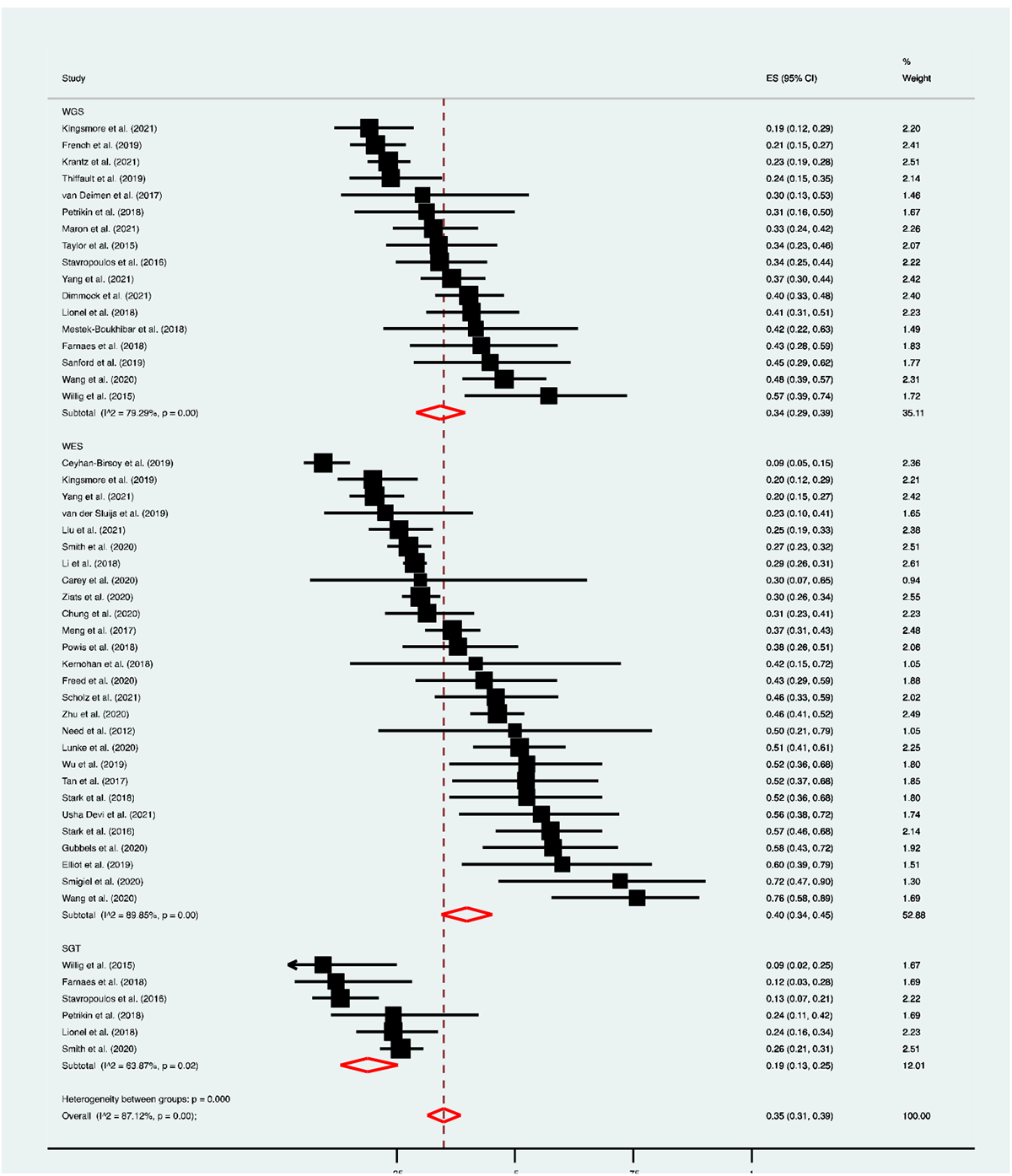
Forest plot of the diagnostic utility of WGS, WES, and standard genetic testing (SGT).

**Figure 3:**
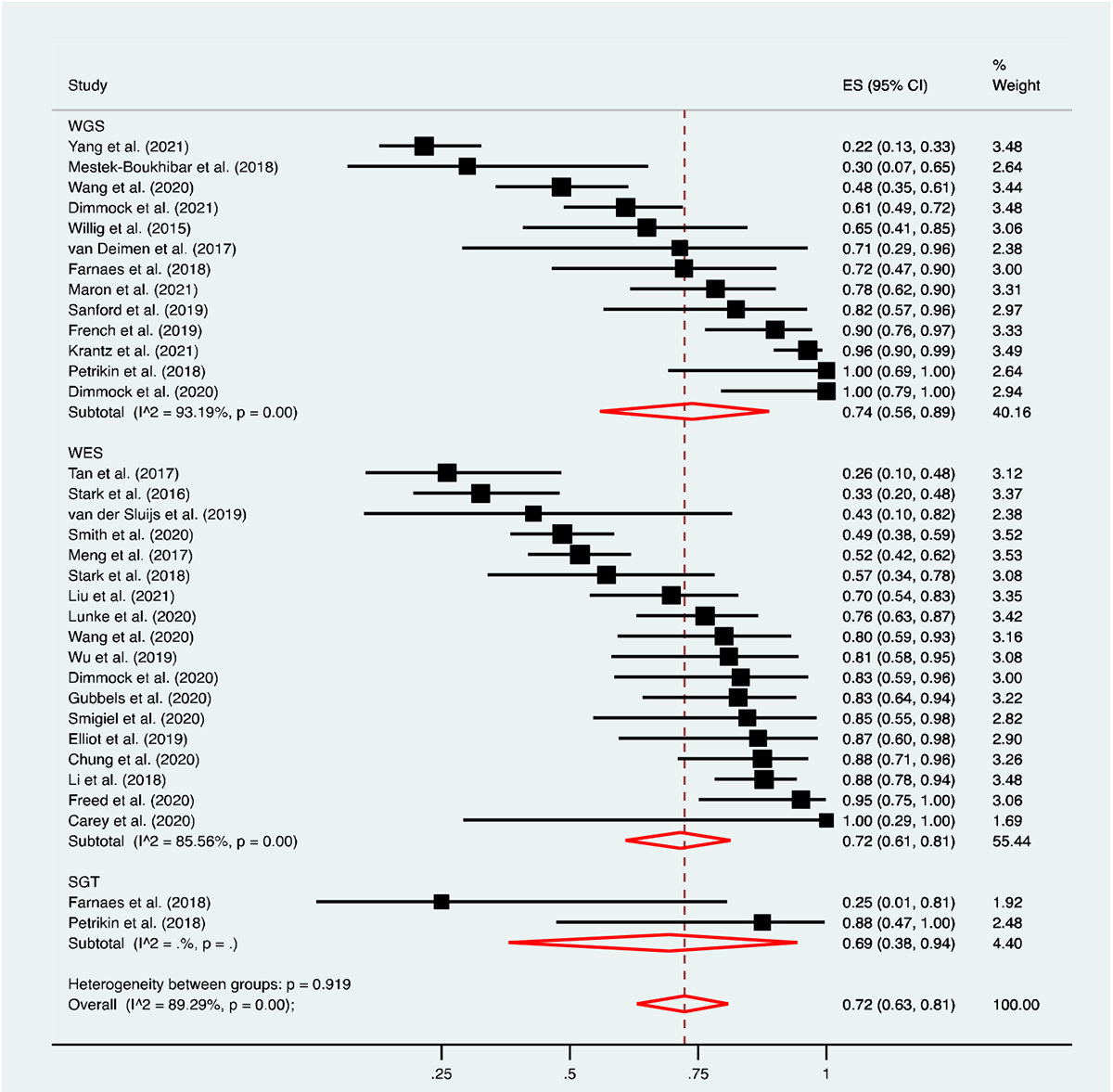
Forest plot of the clinical utility of WGS, WES, and standard genetic testing (SGT).

**Figure 4:**
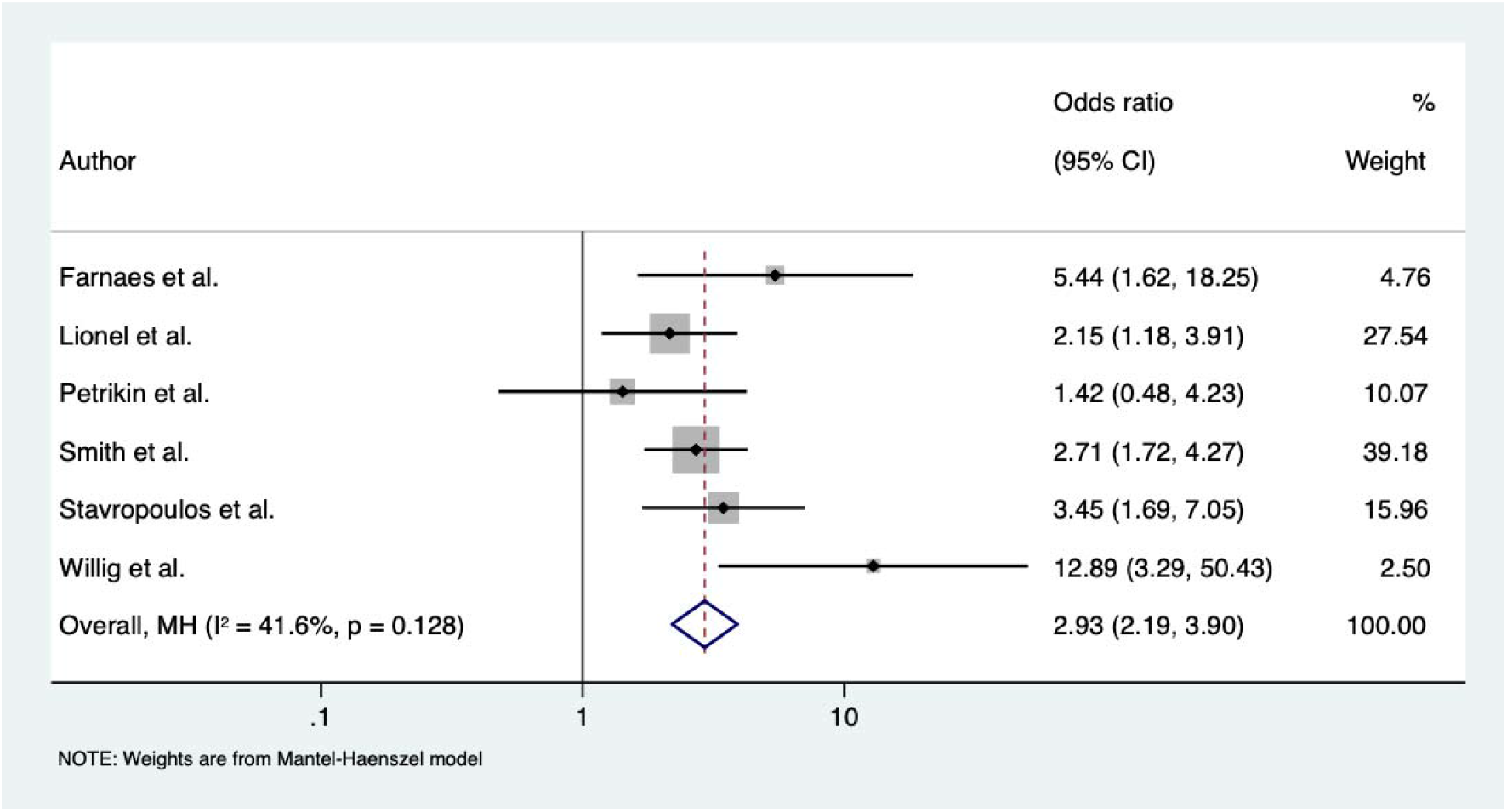
Forest plot of the odds ratio of WGS, WES, and standard genetic testing (SGT).

**Table X:**
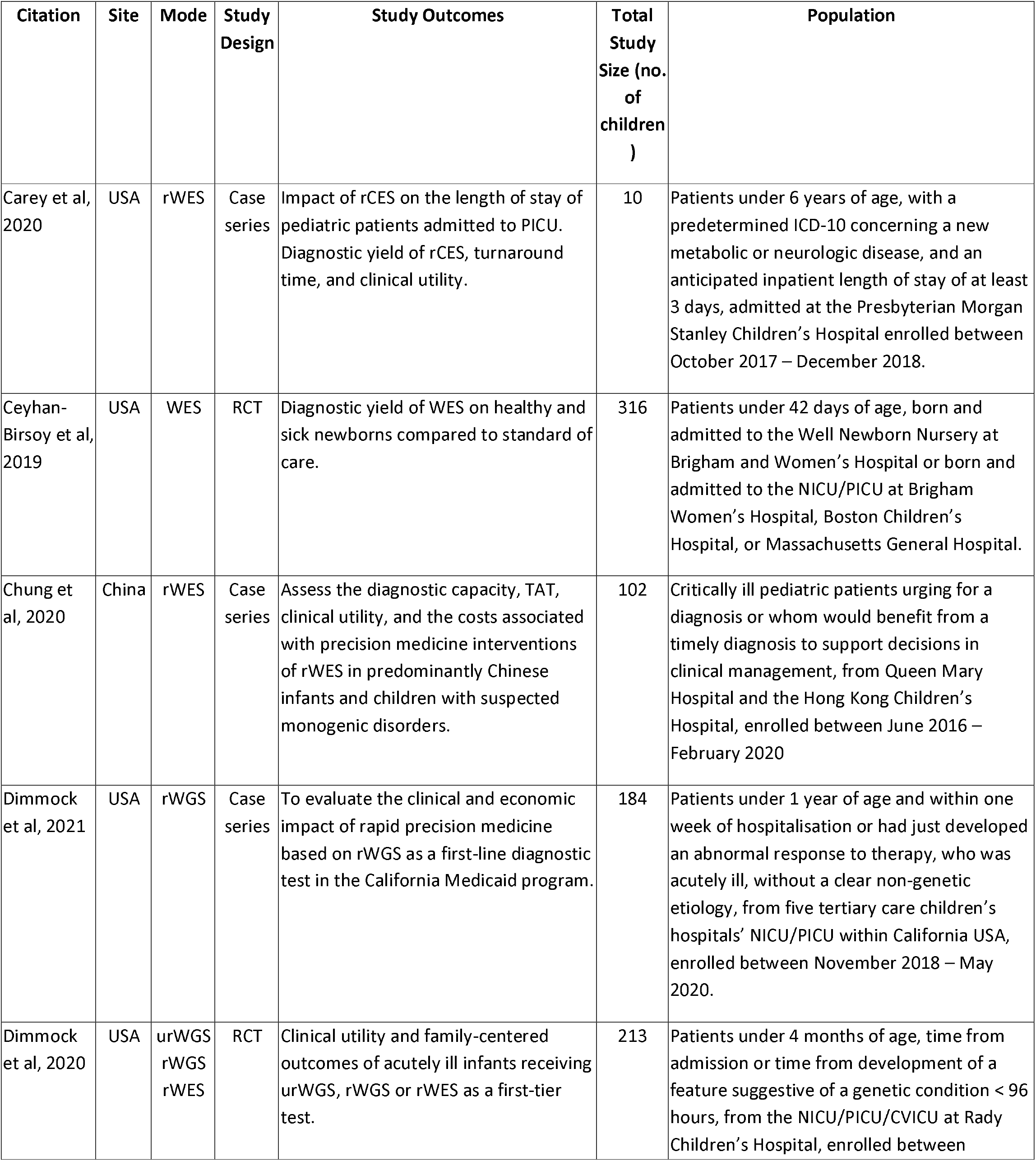
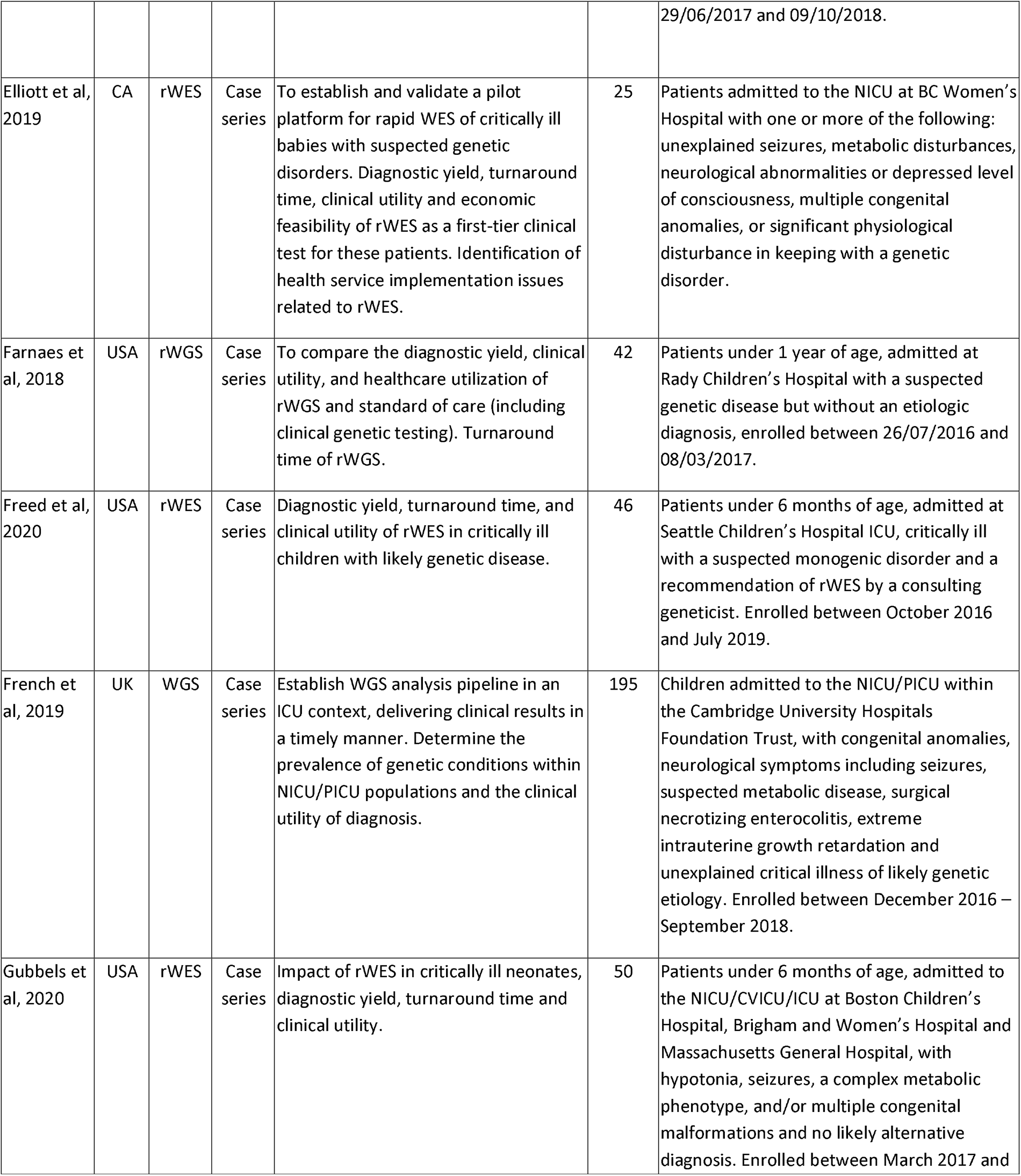
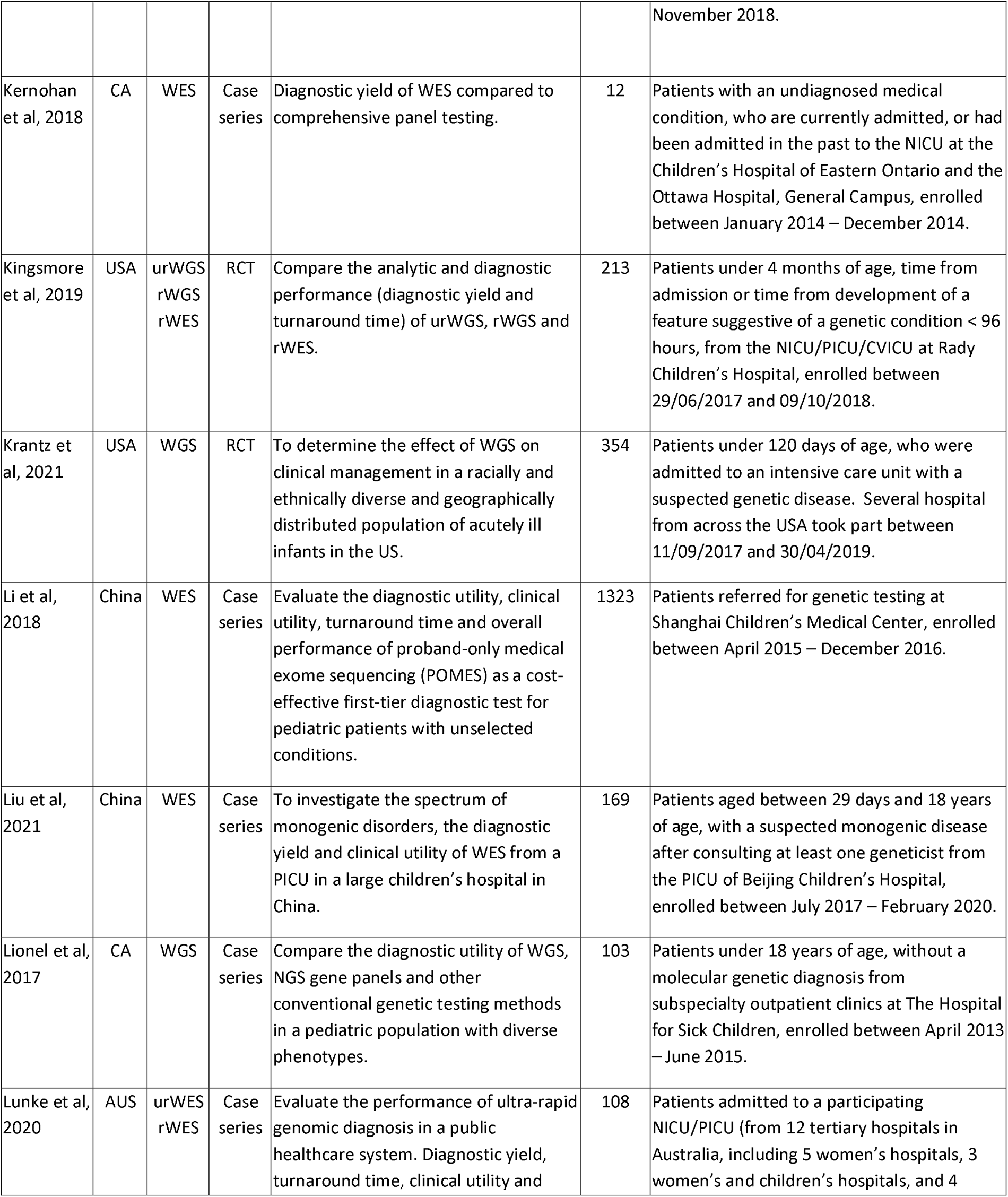
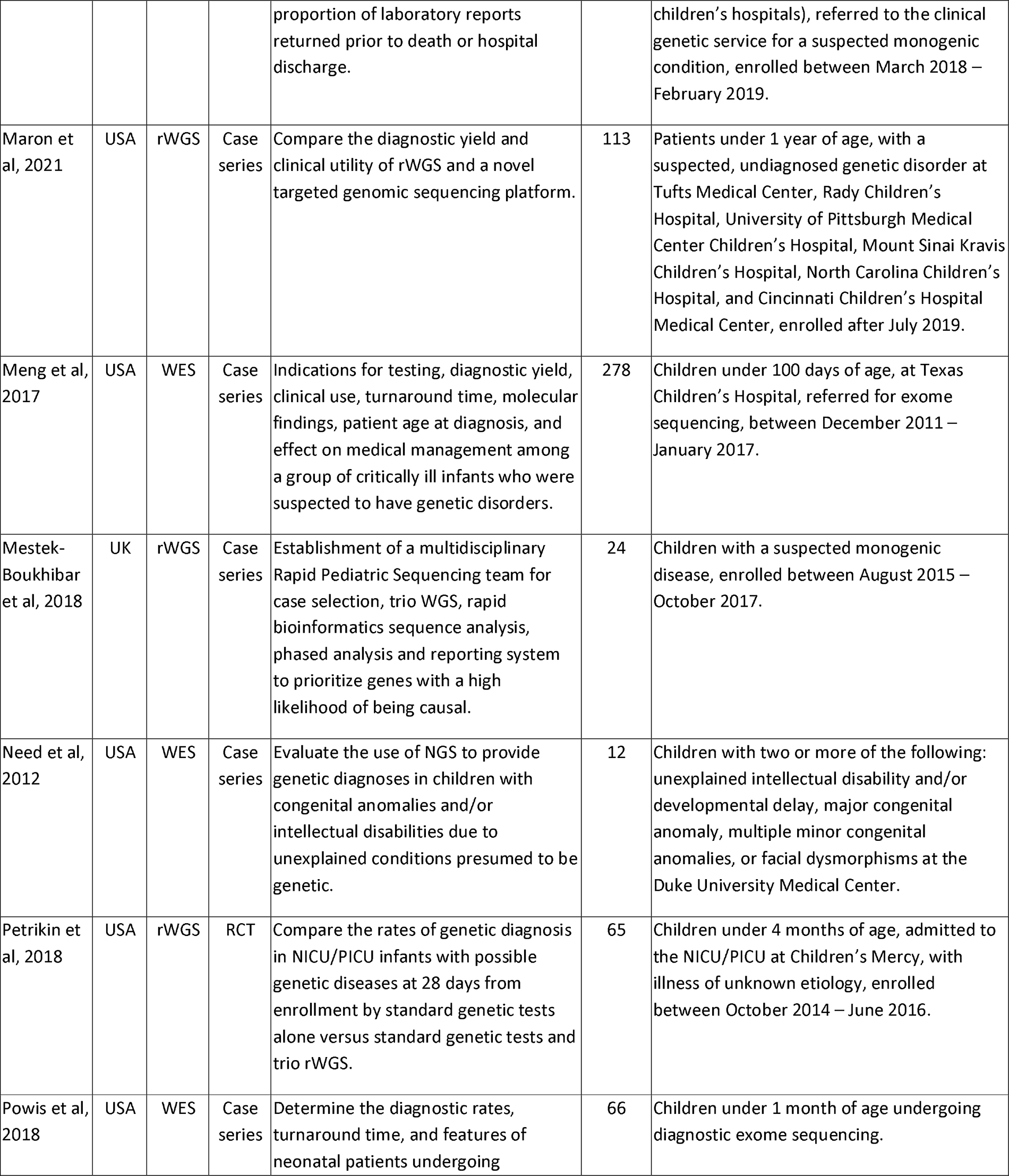
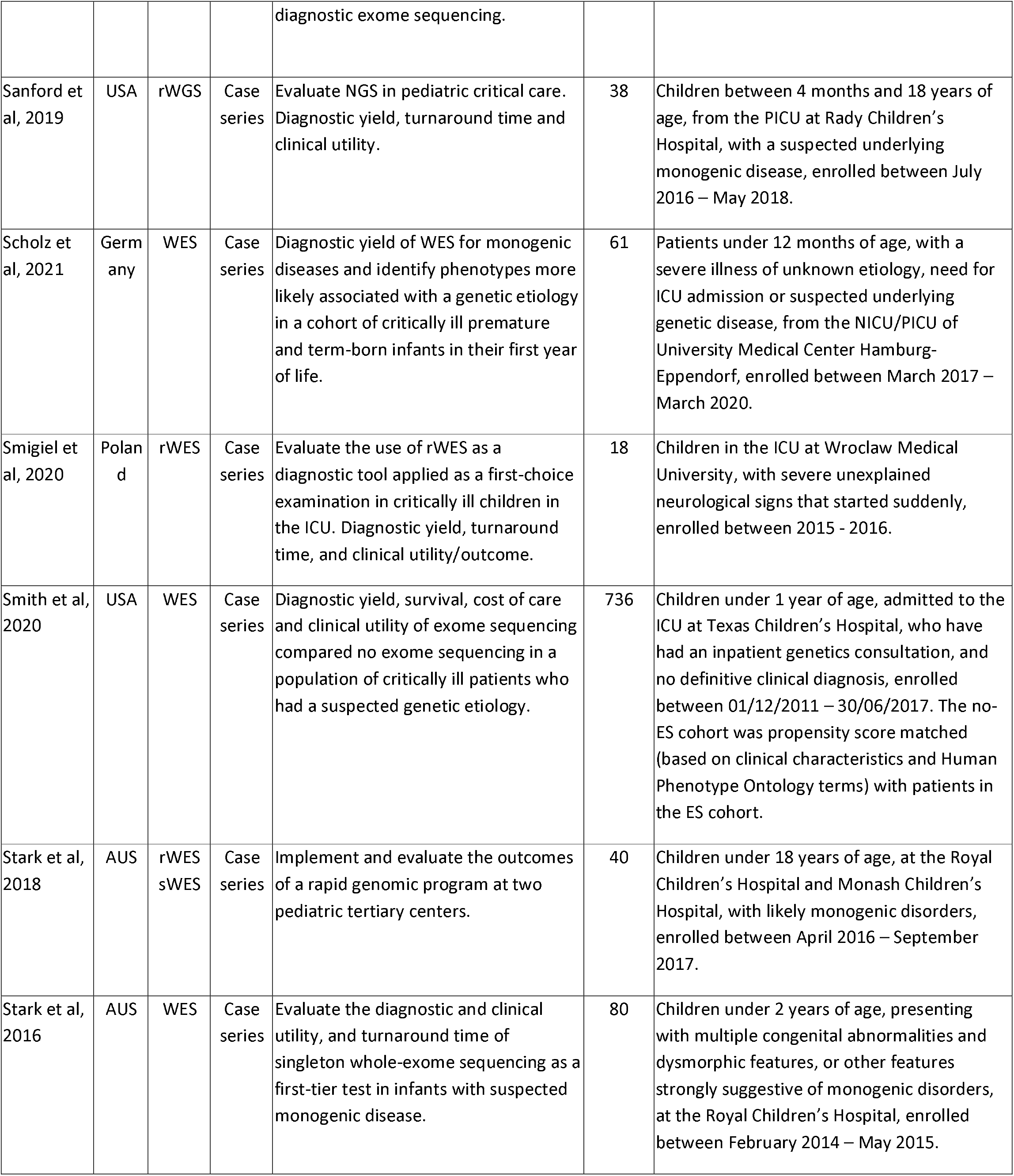
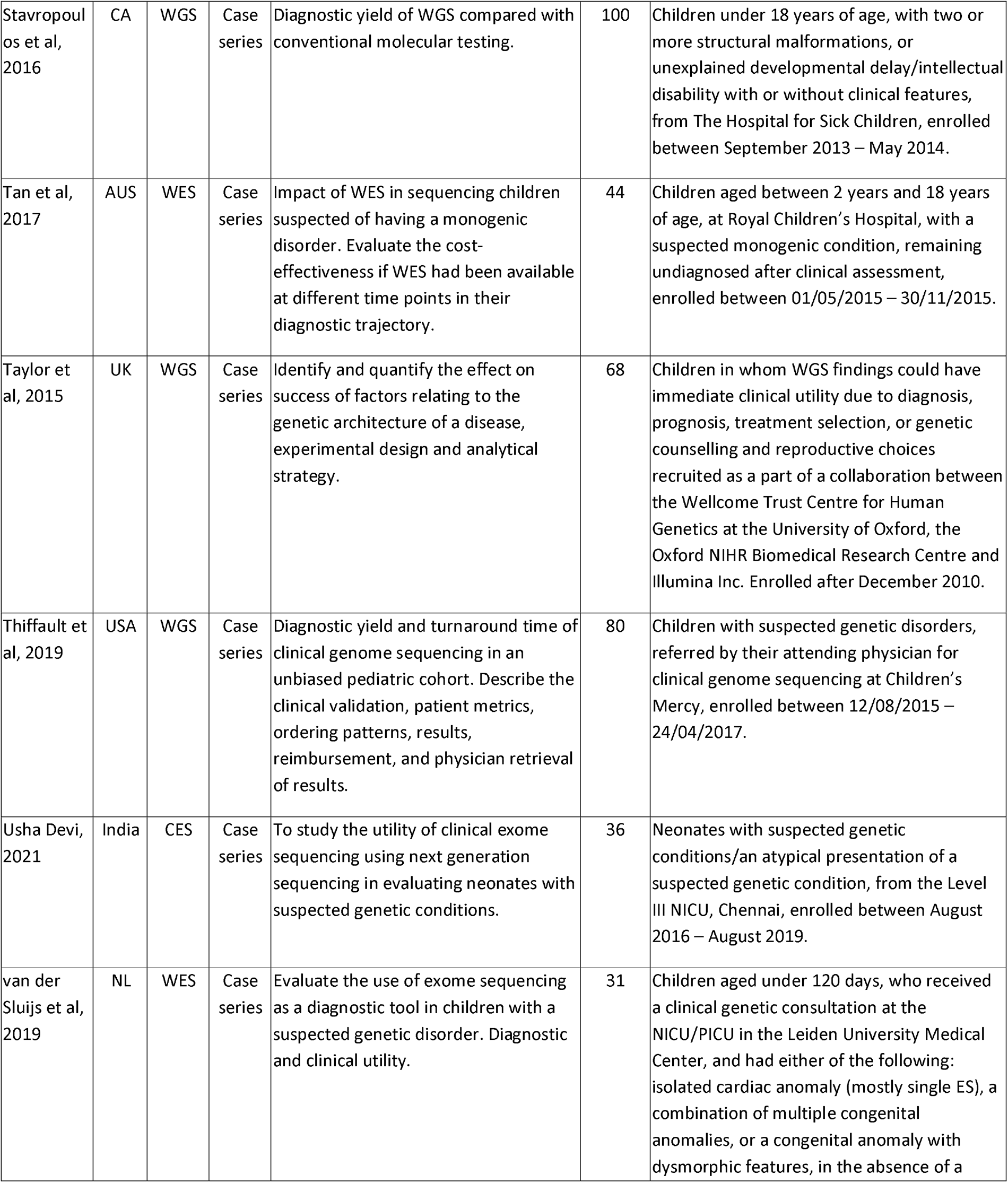
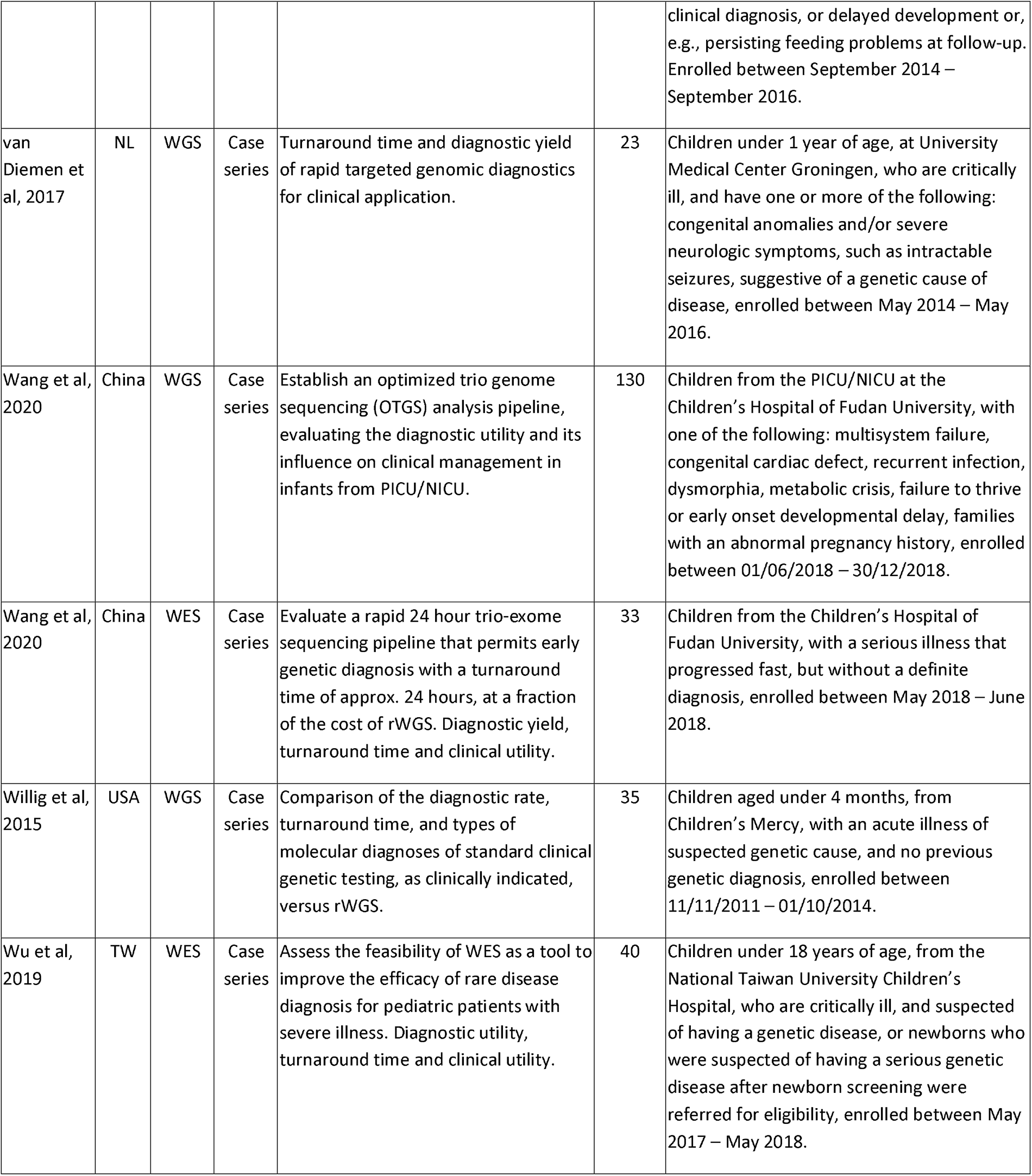
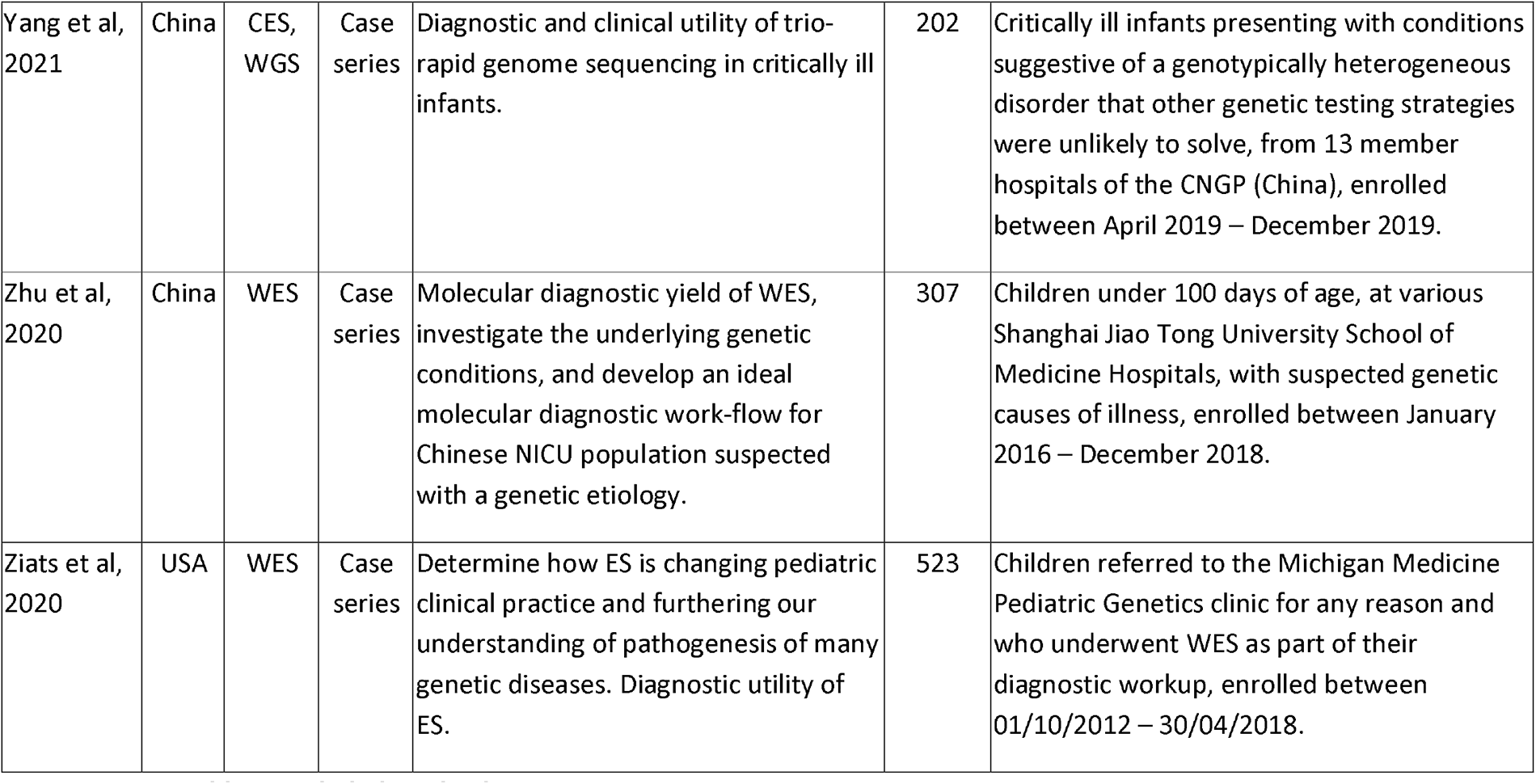
Included Study Characteristics.

### 7.0 Discussion

Since the early 2010’s, WGS and WES have gained recognition for the diagnosis of genetic diseases, however, widespread clinical use and thorough guidelines still do not exist. This systematic review identified thirty-six publications, comprising a total of 5540 children, reporting the diagnostic or clinical utility of WGS and WES. The pooled diagnostic utility showed that WES (0.40, 95% CI 0.34-0.45, *I^2^*=90%), was qualitatively greater than WGS (0.34, 95% CI 0.29-0.39, *I^2^*=79%), and SGT (0.19, 95% CI 0.13-0.25, *I^2^*=64%). Only 6 (14%) included studies reported results of a comparator test, including CMA, Sanger sequencing, single gene tests, and gene panel testing. As such, comparisons of any statistical pooling were highly susceptible to confounding from factors; possible factors included: testing procedures, patient factors, such as consanguinity, eligibility criteria, or clinician input. This was evident within the severe levels of statistical heterogeneity between the study groups. These results suggest that CMA, Sanger sequencing, and other genetic tests, should no longer be considered the best genomic test for the diagnosis of children with suspected genetic disease, in terms of diagnostic utility; rather, WGS and WES should be considered the first-line genomic test.

While diagnostic utility is the primary measure of importance for a clinical diagnostic test, the clinical utility of WGS and WES is of high importance in order to improve the clinical outcomes of children with suspected genetic diseases. Forty-two (98%) of the included papers reported the diagnostic utility of WGS and/or WES, however, only thirty-one (72%) reported the clinical utility after diagnosis. The clinical utility of WGS and WES was measured in numerous ways throughout the studies, including clinician surveys, EHR reviews, or a combination of both. The heterogeneity within the clinical presentations and genetic origins of diseases, and the resulting numerous medical interventions, can result in a number of possible changes in medical management, thus increasing the difficulty of generalising measures of clinical utility. We defined clinical utility as any change in management within infants who have obtained a diagnosis from WGS, WES, or SGT, as determined through clinician survey and/or EHR review. Changes included further testing, transferral to palliative care, and withdrawal of support, but excluded genetic counselling and parental reproductive planning, as genetic counselling should be offered to all children and their families, regardless of diagnostic result, and reproductive planning for parents does not affect the diagnosed child’s clinical status. The pooled clinical utility showed that WGS (0.74, 95% CI 0.56-0.89, *I^2^*=93%), was qualitatively greater than WES (0.72, 95% CI 0.61-0.81, *I^2^*=86%), while both were qualitatively greater than SGT (0.69, 95% CI 0.38-0.94). However, the results showed severe heterogeneity (*I^2^*>75%) for WGS and WES, precluding a statistical comparison. Of the 6 (14%) papers to report results of a comparator test, only 2 (5%) reported outcomes of clinical management, which meant heterogeneity could not be calculated for the SGT group, and comparisons of statistical pooling would not be appropriate. Interestingly, some studies reported the clinical utility for all of the infants enrolled, including non-diagnosed patients, with some clinicians regarding WGS and WES to have a considerable negative predictive value, employing an informal Bayesian inferential reasoning, whereby negative genomic sequencing results revised the posterior probabilities of differential diagnoses. These studies suggest that, even after a non-diagnostic result, WGS and WES have some clinical utility, although changes in management were 10.1-fold more likely when results were positive (95% CI 4.7-22.4).(^31, 35, 46–48, 50, 52, 57^) Changes in management for non-diagnosed participants included cancellation of planned tissue biopsies, cessation of medications, subspecialist referrals, and screening recommendations, typically as a result of non-genetic diagnoses thought to be more likely.

If the overall diagnostic success of WGS and WES was 34% and 40%, respectively, and the overall clinical success of WGS and WES was the 74% and 72%, respectively, showing that WGS and WES were highly beneficial in the treatment, management, and therefore, survival of these children, then there is a strong case for standardising WGS/WES in newborns. Although it could be argued that WGS and WES should be used as the first-line genomic test for children with suspected genetic diseases, rather than for all children as part of the NBS programme, in order to fully utilise the clinical utility of WGS/WES, they should be used as part of the NBS procedure, before any symptoms manifest and the risk of morbidity and mortality increases. Testing approaches of parent-child trios, duos, and singletons varied between papers; although these sub-group approaches were not analysed in this review, the testing of parent-child trios is considered to be superior to singleton and duo testing. This is thought to be due to the ability of trio testing allowing for heritability to be determined, more specifically, whether the variants detected were inherited through the parents or de novo mutations. Of the papers that reported the heritability of variants, de novo variants accounted for 596/1381 (43%) detected by WGS and WES in total. This included P/LP variants that led to diagnoses, variants deemed to be an incidental finding, and VUS. De novo variants are of significant importance, in the context of WGS and WES for all newborns, as preconception genetic screening would not detect these variants.

There were several limitations to this meta-analysis. We were limited to analysing WGS and WES on cohorts of unwell children with suspected genetic diseases. This was not truly representative of the target population WGS and WES would be used for NBS. During initial searches, only one study was identified that researched the use of WGS within healthy and unwell cohorts.(^38^) This paper was included in this study; however, the healthy cohort was omitted from data extraction and analysis due to not meeting the patient selection criteria, presenting a risk of bias, and source of heterogeneity. The field would benefit from further studies on the diagnostic and clinical utility of WGS and WES on an unbiased healthy cohort, to statistically determine if WGS and WES are worthwhile and cost-effective approaches to NBS. The highest level of evidence for clinical interventions is meta-analyses of RCTs (Level 1).(^65^) Our literature search identified only five published RCTs, with two looking at different outcomes from the same trial, while another compared time to receipt of results rather than comparing sequencing methods. Each RCT compared different index tests and reference standards, two looking WES vs WGS, one examining WGS vs SGT, and the other looking et WES vs. SGT. We were, therefore, unable to produce a high-level evidence meta-analysis of WGS and WES compared to SGT. Our review consisted mainly of published studies comprising a Level 2 (non-randomised controlled studies or quasi-experimental studies) and Level 3 evidence (non-experimental descriptive studies, such as comparative studies, correlation studies, and case-control studies). We examined the diagnostic and clinical utility of WGS and WES compared to SGT. However, severe heterogeneity was present across all between-group analyses. This could largely be due to differing rates of consanguinity within the cohort, as well as “cherry-picking” of participants with certain clinical presentations considered to have a high likelihood of a genetic origin. The year of publication could have also played a part; WGS and WES are relatively new techniques whose methodologies and interpretations are expanding with time and further knowledge. The rates of severe heterogeneity could be better explored through a meta-regression to determine the impacts of certain confounding factors on heterogeneity. The meta-analysis did not include the cost-effectiveness of WGS and WES compared to SGT, either in terms of the patient’s diagnostic odyssey or the overall impact on the healthcare system.

### 8.0 Conclusion

In meta-analyses of 43 studies of children with suspected genetic diseases, the diagnostic utility of WES (0.40, 95% CI 0.34-0.45, *I^2^*=90%), was qualitatively greater than WGS (0.34, 95% CI 0.29-0.39, *I^2^*=79%), and SGT (0.19, 95% CI 0.13-0.25, *I^2^*=64%). For the rate of clinical utility, WGS (0.74, 95% CI 0.56-0.89, *I^2^*=93%), was qualitatively greater than WES (0.72, 95% CI 0.61-0.81, *I^2^*=86%), while both were qualitatively greater than SGT (0.69, 95% CI 0.38-0.94). Additional studies are needed to examine the effectiveness of WGS and WES in cohorts of healthy children, particularly RCTs examining the diagnostic and clinical utility, as well as the cost-effectiveness of using these sequencing techniques in this area, in order to truly determine if WGS and WES should become part of the NBS programme.

## Data Availability

All data produced in the present study are available upon reasonable request to the authors

## Abbreviations

ACMG: American College of Medical Genetics
CMA: Chromosomal microarray
EHR: Electronic Health Record
NBS: Newborn screening
NGS: Next-generation sequencing
P/LP: Pathogenic/Likely Pathogenic
SGT: Standard genetic testing
VUS: Variants of unknown significance
WGS: Whole-genome sequencing
WES: Whole-exome sequencing

## PRISMA 2020 Checklist

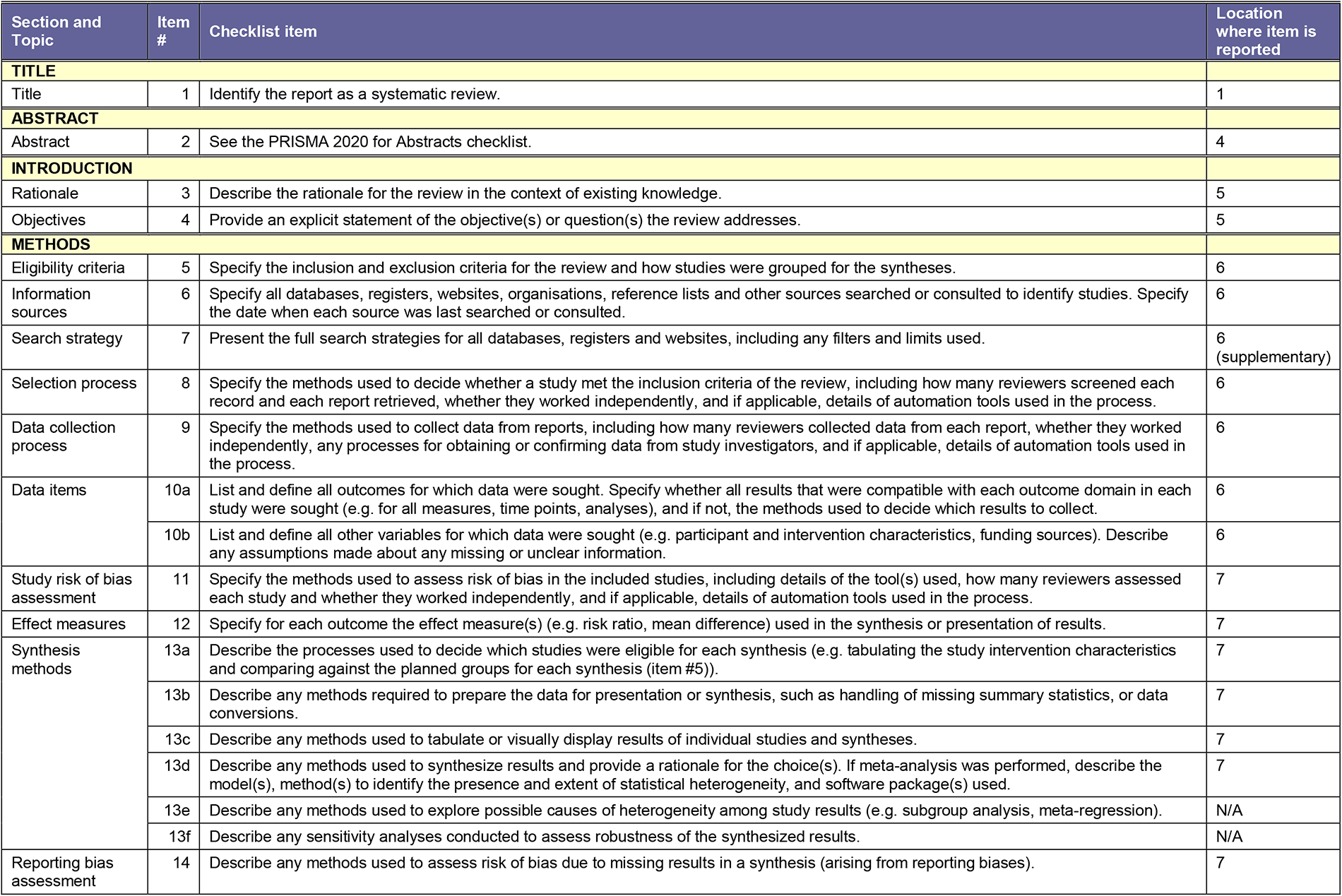
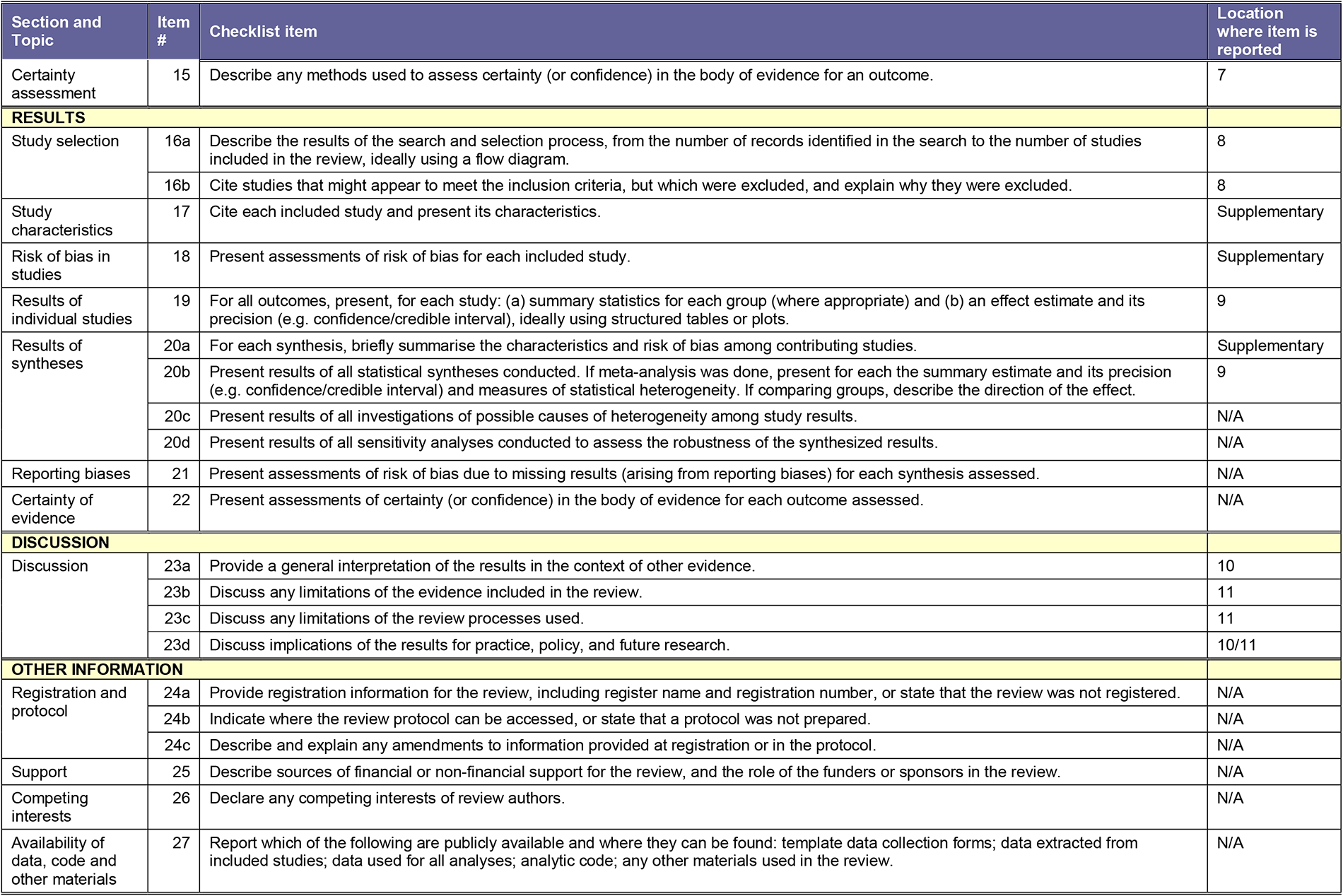

| Section and Topic | Item # | Checklist item | Location where item is reported |
| --- | --- | --- | --- |
| **TITLE** |  |  |  |
| Title | 1 | Identify the report as a systematic review. | 1 |
| **ABSTRACT** |  |  |  |
| Abstract | 2 | See the PRISMA 2020 for Abstracts checklist. | 4 |
| **INTRODUCTION** |  |  |  |
| Rationale | 3 | Describe the rationale for the review in the context of existing knowledge. | 5 |
| Objectives | 4 | Provide an explicit statement of the objective(s) or question(s) the review addresses. | 5 |
| **METHODS** |  |  |  |
| Eligibility criteria | 5 | Specify the inclusion and exclusion criteria for the review and how studies were grouped for the syntheses. | 6 |
| Information sources | 6 | Specify all databases, registers, websites, organisations, reference lists and other sources searched or consulted to identify studies. Specify the date when each source was last searched or consulted. | 6 |
| Search strategy | 7 | Present the full search strategies for all databases, registers and websites, including any filters and limits used. | 6 (supplementary) |
| Selection process | 8 | Specify the methods used to decide whether a study met the inclusion criteria of the review, including how many reviewers screened each record and each report retrieved, whether they worked independently, and if applicable, details of automation tools used in the process. | 6 |
| Data collection process | 9 | Specify the methods used to collect data from reports, including how many reviewers collected data from each report, whether they worked independently, any processes for obtaining or confirming data from study investigators, and if applicable, details of automation tools used in the process. | 6 |
| Data items | 10a | List and define all outcomes for which data were sought. Specify whether all results that were compatible with each outcome domain in each study were sought (e.g. for all measures, time points, analyses), and if not, the methods used to decide which results to collect. | 6 |
|  | 10b | List and define all other variables for which data were sought (e.g. participant and intervention characteristics, funding sources). Describe any assumptions made about any missing or unclear information. | 6 |
| Study risk of bias assessment | 11 | Specify the methods used to assess risk of bias in the included studies, including details of the tool(s) used, how many reviewers assessed each study and whether they worked independently, and if applicable, details of automation tools used in the process. | 7 |
| Effect measures | 12 | Specify for each outcome the effect measure(s) (e.g. risk ratio, mean difference) used in the synthesis or presentation of results. | 7 |
| Synthesis methods | 13a | Describe the processes used to decide which studies were eligible for each synthesis (e.g. tabulating the study intervention characteristics and comparing against the planned groups for each synthesis (item #5)). | 7 |
|  | 13b | Describe any methods required to prepare the data for presentation or synthesis, such as handling of missing summary statistics, or data conversions. | 7 |
|  | 13c | Describe any methods used to tabulate or visually display results of individual studies and syntheses. | 7 |
|  | 13d | Describe any methods used to synthesize results and provide a rationale for the choice(s). If meta-analysis was performed, describe the model(s), method(s) to identify the presence and extent of statistical heterogeneity, and software package(s) used. | 7 |
|  | 13e | Describe any methods used to explore possible causes of heterogeneity among study results (e.g. subgroup analysis, meta-regression). | N/A |
|  | 13f | Describe any sensitivity analyses conducted to assess robustness of the synthesized results. | N/A |
| Reporting bias assessment | 14 | Describe any methods used to assess risk of bias due to missing results in a synthesis (arising from reporting biases). | 7 |

PRISMA 2020 Checklist
| Section and Topic | Item # | Checklist item | Location where item is reported |
| --- | --- | --- | --- |
| Certainty assessment | 15 | Describe any methods used to assess certainty (or confidence) in the body of evidence for an outcome. | 7 |
| **RESULTS** |  |  |  |
| Study selection | 16a | Describe the results of the search and selection process, from the number of records identified in the search to the number of studies included in the review, ideally using a flow diagram. | 8 |
|  | 16b | Cite studies that might appear to meet the inclusion criteria, but which were excluded, and explain why they were excluded. | 8 |
| Study characteristics | 17 | Cite each included study and present its characteristics. | Supplementary |
| Risk of bias in studies | 18 | Present assessments of risk of bias for each included study. | Supplementary |
| Results of individual studies | 19 | For all outcomes, present, for each study: (a) summary statistics for each group (where appropriate) and (b) an effect estimate and its precision (e.g. confidence/credible interval), ideally using structured tables or plots. | 9 |
| Results of syntheses | 20a | For each synthesis, briefly summarise the characteristics and risk of bias among contributing studies. | Supplementary |
|  | 20b | Present results of all statistical syntheses conducted. If meta-analysis was done, present for each the summary estimate and its precision (e.g. confidence/credible interval) and measures of statistical heterogeneity. If comparing groups, describe the direction of the effect. | 9 |
|  | 20c | Present results of all investigations of possible causes of heterogeneity among study results. | N/A |
|  | 20d | Present results of all sensitivity analyses conducted to assess the robustness of the synthesized results. | N/A |
| Reporting biases | 21 | Present assessments of risk of bias due to missing results (arising from reporting biases) for each synthesis assessed. | N/A |
| Certainty of evidence | 22 | Present assessments of certainty (or confidence) in the body of evidence for each outcome assessed. | N/A |
| **DISCUSSION** |  |  |  |
| Discussion | 23a | Provide a general interpretation of the results in the context of other evidence. | 10 |
|  | 23b | Discuss any limitations of the evidence included in the review. | 11 |
|  | 23c | Discuss any limitations of the review processes used. | 11 |
|  | 23d | Discuss implications of the results for practice, policy, and future research. | 10/11 |
| **OTHER INFORMATION** |  |  |  |
| Registration and protocol | 24a | Provide registration information for the review, including register name and registration number, or state that the review was not registered. | N/A |
|  | 24b | Indicate where the review protocol can be accessed, or state that a protocol was not prepared. | N/A |
|  | 24c | Describe and explain any amendments to information provided at registration or in the protocol. | N/A |
| Support | 25 | Describe sources of financial or non-financial support for the review, and the role of the funders or sponsors in the review. | N/A |
| Competing interests | 26 | Declare any competing interests of review authors. | N/A |
| Availability of data, code and other materials | 27 | Report which of the following are publicly available and where they can be found: template data collection forms; data extracted from included studies; data used for all analyses; analytic code; any other materials used in the review. | N/A |

From: Page MJ, McKenzie JE, Bossuyt PM, Boutron I, Hoffmann TC, Mulrow CD, et al. The PRISMA 2020 statement: an updated guideline for reporting systematic reviews. BMJ 2021; 372: n71. doi: 10.1136/bmj.n71

For more information, visit: http://www.prisma-statement.org/

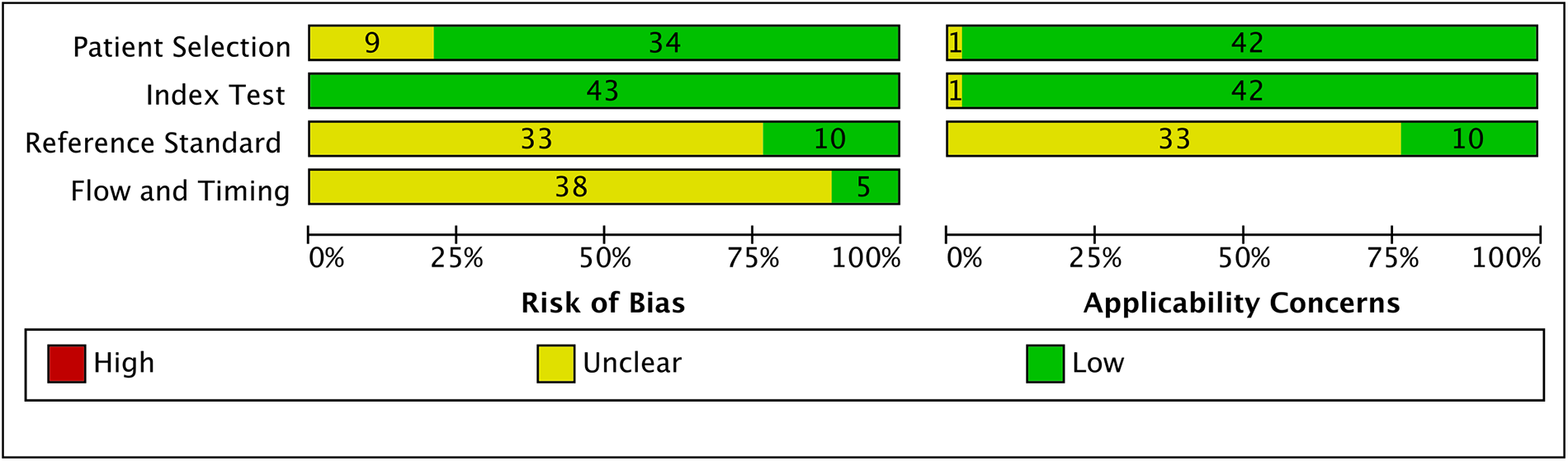

**P:** seriously ill children, infants, newborns with suspected genetic disorders

**I:** whole exome sequencing, whole genome sequencing

**C:** single-gene testing/standard newborn screening

**O:** genetic/clinical diagnosis, time to diagnosis, change in treatment/management

**Concept 1:** children, infant, newborn

- MeSH terms: “Infant”[Mesh] OR “Infant, Newborn”[Mesh] OR “Child”[Mesh]
- Key search terms: child*[tw] OR infant[tw] OR newborn[tw]

**Concept 2:** whole genome sequencing, whole exome sequencing

- MeSH terms: “Whole Genome Sequencing”[Mesh] OR “Whole Exome Sequencing”[Mesh] OR “Genetic Testing”[Mesh] OR “High-Throughput Nucleotide Sequencing”[Mesh]
- Key search terms: WGS[tw] OR rWGS[tw] OR urWGS[tw] OR WES[tw] OR rWES[tw] OR “whole genome seqeunc*”[tw] OR “whole exome sequenc*”[tw]

**Concept 3:** critically ill, ICU, NICU, PICU

- MeSH terms: “Critical Illness”[Mesh] OR “Intensive Care Units”[Mesh] OR “Intensive Care, Neonatal”[Mesh] OR “Intensive Care Units, Pediatric”[Mesh] OR “Intensive Care Units, Neonatal”[Mesh] OR “Critical Care”[Mesh]
- Key search terms: “critically ill”[tw] OR ICU[tw] OR NICU[tw] OR PICU[tw]

### PubMed Search

#1 “Infant”[Mesh] OR “Infant, Newborn”[Mesh] OR “Child”[Mesh] OR child*[tw] OR infant[tw] OR newborn[tw] OR baby[tw] OR neonate[tw]

#2 “Whole Genome Sequencing”[Mesh] OR “Whole Exome Sequencing”[Mesh] OR “Genetic Testing”[Mesh] OR “High-Throughput Nucleotide Sequencing”[Mesh] OR WGS[tw] OR rWGS[tw] OR urWGS[tw] OR WES[tw] OR rWES[tw] OR “whole genome seqeunc*”[tw] OR “whole exome sequenc*”[tw] OR “whole-genome sequenc*”[tw] OR “whole-exome sequenc*”[tw] OR “genomic sequenc*”[tw] OR “exome sequenc*”[tw] OR “genome sequenc*”[tw] OR TES[tw] OR TGS[tw] OR miseq[tw] OR hiseq[tw] OR “ion torrent”[tw] OR “clinical exome sequenc*”[tw] OR CES[tw]

#3 “Critical Illness”[Mesh] OR “Intensive Care Units”[Mesh] OR “Intensive Care, Neonatal”[Mesh] OR “Intensive Care Units, Pediatric”[Mesh] OR “Intensive Care Units, Neonatal”[Mesh] OR “Critical Care”[Mesh] OR “critically ill”[tw] “critical illness”[tw] OR “gravely ill”[tw] OR “severely ill”[tw] OR ill[tw] OR unwell[tw] OR sick[tw] OR ICU[tw] OR NICU[tw] OR PICU[tw]

#4 #1 AND #2 AND #3

### Ovid MEDLINE® and Epub Ahead of Print, In-Process, In-Data-Review & Other Non-Indexed Citations and Daily 1946 to October Week 3 2021

**#1** exp whole genome sequencing/

**#2** exp whole exome sequencing/

**#3** “whole genome sequenc*”.ti,ab,mp.

**#4** “whole-genome sequenc*”.ti,ab,mp.

**#5** “whole exome sequenc*”.ti,ab,mp.

**#6** “whole-exome sequenc*”.ti,ab,mp.

**#7** “genomic sequenc*”.ti,ab,mp.

**#8** wgs.ti,ab,mp.

**#9** wes.ti,ab,mp.

**#10** rwgs.ti,ab,mp.

**#11** urwgs.ti,ab,mp.

**#12** rwes.ti,ab,mp.

**#13** “exome sequenc*”.ti,ab,mp.

**#14** “genome sequenc*”.ti,ab,mp.

**#15** tes.ti,ab,mp.

**#16** tgs.ti,ab,mp.

**#17** miseq.ti,ab,mp.

**#18** hiseq.ti,ab,mp.

**#19** “ion torrent”.ti,ab,mp.

**#20** “clinical exome sequenc*”.ti,ab,mp.

**#21** ces.ti,ab,mp.

**#22** 1 or 2 or 3 or 4 or 5 or 6 or 7 or 8 or 9 or 10 or 11 or 12 or 13 or 14 or 15 or 16 or 17 or 18 or 19 or 20 or 21

**#23** exp child/

**#24** exp infant/

**#25** exp newborn/

**#26** child*.ti,ab,mp.

**#27** infant.ti,ab,mp.

**#28** newborn.ti,ab,mp.

**#29** baby.ti,ab,mp.

**#30** neonate.ti,ab,mp.

**#31** 23 or 24 or 25 or 26 or 27 or 28 or 29 or 30

**#32** exp Critical Illness/

**#33** exp intensive care unit/

**#34** exp intensive care units, neonatal/

**#35** exp pediatric intensive care unit/

**#36** exp intensive care/

**#37** “critical illness”.ti,ab,mp.

**#38** “critically ill”.ti,ab,mp.

**#39** “gravely ill”.ti,ab,mp.

**#40** “severely ill”.ti,ab,mp.

**#41** ill.ti,ab,mp.

**#42** unwell.ti,ab,mp.

**#43** sick.ti,ab,mp. #44 exp critical care/

**#45** icu.ti,ab,mp.

**#46** nicu.ti,ab,mp.

**#47** picu.ti,ab,mp.

**#48** 32 or 33 or 34 or 35 or 36 or 37 or 38 or 39 or 40 or 41 or 42 or 43 or 44 or 45 or 46 or 47

**#49** 22 and 31 and 48

### Scopus Search

**#1** TITLE-ABS-KEY(child* OR infant OR newborn OR baby OR neonate)

**#2** TITLE-ABS-KEY(“whole genome sequenc*” OR “whole-genome sequenc*” OR “whole exome sequenc*” OR “whole-exome sequenc*” OR “genomic sequenc*” OR {WGS} OR {rWGS} OR {urWGS} OR {WES} OR {rWES} OR “exome sequenc*” OR “genome sequenc*” OR {TES} OR {TGS} OR {miseq} OR {hiseq} OR “ion torrent” OR “clinical exome sequenc*” OR {CES})

**#3** TITLE-ABS-KEY(“critically ill” OR “critical illness” OR “gravely ill” OR “severely ill” OR ill OR unwell OR sick OR ICU OR NICU OR PICU)

**#4** #1 AND #2 AND #3

### Web of Science Search

**#1** ts=child*

**#2** ts=infant

**#3** ts=newborn

**#4** ts=baby

**#5** ts=neonate

**#6** #1 OR #2 OR #3 OR #4 OR #5

**#7** ts=“whole genome sequenc*”

**#8** ts=“whole-genome sequenc*”

**#9** ts=“whole exome sequenc*”

**#10** ts=“whole-exome sequenc*”

**#11** ts=“genomic sequenc*”

**#12** ts=WGS

**#13** ts=rWGS

**#14** ts=urWGS

**#15** ts=WES

**#16** ts=rWES

**#17** ts=“exome sequenc*”

**#18** ts=“genome sequenc*”

**#19** ts=TES

**#20** ts=TGS

**#21** ts=miseq

**#22** ts=hiseq

**#23** ts=“ion torrent”

**#24** ts=“clinical exome sequenc*”

**#25** ts=CES

**#26** #7 OR #8 OR #9 OR #10 OR #11 OR #12 OR #13 OR #14 OR #15 OR #16 OR #17 OR #18 OR #19 OR #20 OR #21 OR #22 OR #23 OR #24 OR #25

**#27** ts=“critically ill”

**#28** ts=“critical illness”

**#29** ts=“gravely ill”

**#30** ts=“severely ill”

**#31** ts=ill

**#32** ts=unwell

**#33** ts=sick

**#34** ts=ICU

**#35** ts=NICU

**#36** ts=PICU

**#37** #27 OR #28 OR #29 OR #30 OR #31 OR #32 OR #33 OR #34 OR #35 OR #36

**#38** #6 AND #26 AND #37

### EMBASE search 1980 to 2021 Week 41

**#1** exp whole genome sequencing/

**#2** exp whole exome sequencing/

**#3** “whole genome sequenc*”.ti,ab,mp.

**#4** “whole-genome sequenc*”.ti,ab,mp.

**#5** “whole exome sequenc*”.ti,ab,mp.

**#6** “whole-exome sequenc*”.ti,ab,mp.

**#7** “genomic sequenc*”.ti,ab,mp.

**#8** wgs.ti,ab,mp.

**#9** wes.ti,ab,mp.

**#10** rwgs.ti,ab,mp.

**#11** urwgs.ti,ab,mp.

**#12** rwes.ti,ab,mp.

**#13** “exome sequenc*”.ti,ab,mp.

**#14** “genome sequenc*”.ti,ab,mp.

**#15** tes.ti,ab,mp.

**#16** tgs.ti,ab,mp.

**#17** miseq.ti,ab,mp.

**#18** hiseq.ti,ab,mp.

**#19** “ion torrent”.ti,ab,mp.

**#20** “clinical exome sequenc*”.ti,ab,mp.

**#21** ces.ti,ab,mp.

**#22** 1 or 2 or 3 or 4 or 5 or 6 or 7 or 8 or 9 or 10 or 11 or 12 or 13 or 14 or 15 or 16 or 17 or 18 or 19 or 20 or 21

**#23** exp child/

**#24** exp infant/

**#25** exp newborn/

**#26** child*.ti,ab,mp.

**#27** infant.ti,ab,mp.

**#28** newborn.ti,ab,mp.

**#29** baby.ti,ab,mp.

**#30** neonate.ti,ab,mp.

**#31** 23 or 24 or 25 or 26 or 27 or 28 or 29 or 30

**#32** exp Critical Illness/

**#33** exp intensive care unit/

**#34** exp newborn intensive care/

**#35** exp pediatric intensive care unit/

**#36** exp intensive care/

**#37** “critical illness”.ti,ab,mp.

**#38** “critically ill”.ti,ab,mp.

**#39** “gravely ill”.ti,ab,mp.

**#40** “severely ill”.ti,ab,mp.

**#41** exp critically ill patient/

**#42** ill.ti,ab,mp.

**#43** unwell.ti,ab,mp.

**#44** sick.ti,ab,mp.

**#45** icu.ti,ab,mp.

**#46** nicu.ti,ab,mp.

**#47** picu.ti,ab,mp.

**#48** 32 or 33 or 34 or 35 or 36 or 37 or 38 or 39 or 40 or 41 or 42 or 43 or 44 or 45 or 46 or 47

**#49** 22 and 31 and 48

### Cochrane library – Cochrane Central Register of Controlled Trials (Issue 10 of 12, October 2021)

**#1** MeSH descriptor: [Infant] explode all trees

**#2** MeSH descriptor: [Infant, Newborn] explode all trees

**#3** MeSH descriptor: [Child] explode all trees

**#4** child*

**#5** infant

**#6** newborn

**#7** baby

**#8** neonate

**#9** #1 OR #2 OR #3 OR #4 OR #5 OR #6 OR #7 OR #8

**#10** MeSH descriptor: [Whole Genome Sequencing] explode all trees

**#11** MeSH descriptor: [Whole Exome Sequencing] explode all trees

**#12** MeSH descriptor: [Genetic Testing] explode all trees

**#13** MeSH descriptor: [High-Throughput Nucleotide Sequencing] explode all trees

**#14** WGS

**#15** rWGS

**#16** urWGS

**#17** WES

**#18** rWES

**#19** “whole genome sequenc*”

**#20** “whole-genome sequenc*”

**#21** “whole exome sequenc*”

**#22** “whole-exome sequenc*”

**#23** “genomic sequenc*”

**#24** “exome sequenc*”

**#25** “genome sequenc*”

**#26** TES

**#27** TGS

**#28** miseq

**#29** hiseq

**#30** “ion torrent”

**#31** “clinical exome sequenc*”

**#32** CES

**#33** #10 OR #11 OR #12 OR #13 OR #14 OR #15 OR #16 OR #17 OR #18 OR #19 OR #20 OR #21 OR #22 OR #23 OR #24 OR #25 OR #26 OR #27 OR #28 OR #29 OR #30 OR #31 OR #32

**#34** MeSH descriptor: [Critical Illness] explode all trees

**#35** MeSH descriptor: [Intensive Care Units] explode all trees

**#36** MeSH descriptor: [Intensive Care Units, Neonatal] explode all trees

**#37** MeSH descriptor: [Intensive Care Units, Pediatric] explode all trees

**#38** MeSH descriptor: [Critical Care] explode all trees

**#39** “critically ill”

**#40** “critical illness”

**#41** “gravely ill”

**#42** “severely ill”

**#43** ill

**#44** unwell

**#45** sick

**#46** ICU

**#47** NICU

**#48** PICU

**#49** #34 OR #35 OR #36 OR #37 OR #38 OR #39 OR #40 OR #41 OR #42 OR #43 OR #44 OR #45 OR #46 OR #47 OR #48

**#50** #9 AND #33 AND #49

## Notes

### Competing Interest Statement

The authors have declared no competing interest.

### Funding Statement

This study did not receive any funding

